# Sleep Health During Recovery After Stroke

**DOI:** 10.1101/2025.04.13.25325734

**Authors:** George D. Fulk, Karen J. Klingman, Sandra A. Billinger, Bria L. Bartsch, Amanda J. Britton-Carpenter, Keenan Batts, Mary Forgea, Pamela W. Duncan, Emily Peterson

## Abstract

**Study Objectives:** People with stroke are susceptible to developing sleep disorders, which may negatively impact recovery. Little is known about sleep health (SH) broadly and its impact on recovery after stroke. The purpose of this study was to explore factors that are associated with SH during recovery after stroke.

**Methods:** Data were collected from 90 participants at 10-, 60-, and 90-days post stroke without moderate to severe obstructive sleep apnea. Sleep health was measured by combining data from actigraphy and self-report to create an SH score that reflected regularity, satisfaction, alertness, timing, efficiency, and duration of sleep. Factors that may be associated with SH that were collected were Patient Health Questionnaire 9 (PHQ-9), Montreal Cognitive Assessment (MOCA), Barthel Index (BI), gait speed (GS), oxygen desaturation index (ODI), age, and sex. A cumulative link mixed model (CLMM) was used to determine the association between SH and the independent factors.

**Results:** The CLMM found that the PHQ-9 had a significant negative estimate of-0.116 (p = 0.0002) and that the BI had a significant estimate of 0.016 (p = 0.044). Indicating that higher depressive symptoms and lower functional ability are associated with poorer SH. Time post stroke was not associated with SH

**Discussion:** We identified depression and functional ability as significant determinants of SH. Participants exhibited poor SH while in the hospital and it remained unchanged at 60 and 90 days after stroke. Further evaluation of the likely bidirectional relationship between depression/functional impairment and SH after stroke may lead to targets to improve stroke recovery and SH.

## Introduction

People with stroke may be particularly susceptible to poor sleep and developing sleep disorders. The prevalence of obstructive sleep apnea (OSA) in people with stroke may be as high as 71%,^1^ and the prevalence of insomnia is approximately 38%.^2^ Individuals with stroke who have a sleep disorder are likely to have worse functional outcomes and greater disability than those with stroke without a sleep disorder,^3^ and poor self-reported sleep quality is associated with higher levels of depression.^4–7^ Silva and colleagues reported that depression and functional status explained approximately 30% of the variance in self-reported sleep quality in people with stroke.^6^ Iddagoda and colleagues found that people post stroke who reported poor sleep quality had worse outcomes.^7^ Physical activity also appears to influence sleep in people who have experienced a stroke. During inpatient rehabilitation after a stroke, greater amounts of physical activity are associated with better self-reported sleep, while sedentary time appears to have the opposite effect.^8^ Specific sleep parameters are also associated with poorer outcomes after stroke. Total sleep time of less than six hours/night and lower sleep efficiency is associated with poorer functional outcomes in people with stroke.^9^ People with mild stroke with shorter total sleep time are at higher risk of cognitive impairment.^10^

Considering sleep from a broad perspective, rather than focusing solely on specific sleep disorders, is essential for understanding its impact on health and well-being.^11^ Healthy sleep facilitates improved mental, physical, and social health. The American Heart Association included sleep as one of eight essential health behaviors and factors that are important for cardiovascular health.^12^ Different aspects of sleep, both objective and subjective components, encompass sleep health. Buysse^11^ describes six aspects of sleep that comprise sleep health (SH): regularity, satisfaction, alertness/sleepiness, timing, efficiency, and duration. Along with nutrition and physical, sleep is an important part of a general healthy lifestyle.

There is growing evidence that sleep disorders, self-reported sleep quality, and certain sleep parameters play an important role in stroke recovery. A broader view of sleep that encompasses sleep health has not been used in people with stroke. A better understanding of the factors that influence SH may provide guidance in developing targeted interventions to improve recovery after stroke. The purpose of this study was to explore factors that are associated with SH during recovery after stroke in people without moderate to severe OSA. Additionally, we sought to explore how sleep health evolves during recovery after stroke. We hypothesized that depression, functional ability, cognition, and walking capability would be significantly associated with sleep health; and that sleep health would improve after discharge the hospital.

## Methods

Participants were recruited from two inpatient rehabilitation hospitals, one in the Northeast and one in the Midwest of the United States, and are part of a larger, ongoing cohort study to determine the prevalence of non-obstructive sleep apnea (OSA) sleep disorders and their impact on recovery after stroke.^13^ Inclusion criteria were diagnosis of stroke, >= 18 years old, National Institutes of Health Stroke Scale (NIHSS) item 1a score < 2 (alert or not alert but arousable by minor stimulation to obey, answer, or respond), and able to provide informed consent or assent, with the participant’s legal guardian providing consent. Exclusion criteria were pre-stroke diagnosis of OSA, oxygen desaturation index (ODI) >= 15 that was taken at 10 days post stroke, living in nursing home or assisted living prior to stroke, unable to ambulate at least 150’ independently prior to stroke, other neurologic health condition that could impact recovery, pregnancy, recent hemicraniectomy or suboccipital craniectomy, planned discharge > 150 miles from recruiting hospital, and global aphasia as determined by a score of 3 on NIHSS item 9. All participants provided verbal consent or assent (with participant’s legal guardian providing consent), and the study was approved by all local Institutional Review Boards (IRBs) and a central IRB. People with pre-existing OSA or ODI >=15 were excluded because the purpose of the parent study was to determine the prevalence of non-obstructive sleep apnea (OSA) sleep disorders and their impact on recovery after stroke.^13^

For these analyses, the following data collected at 10-, 60-, and 90-days post stroke were used: wrist worn actigraphy to measure sleep parameters, Insomnia Severity Index (ISI),^14,15^ Epworth Sleepiness Scale (ESS),^16,17^ ODI measured while participants wore a Nonin WristOx2 3150 overnight, Patient Health Questionnaire 9 (PHQ-9),^18^ Montreal Cognitive Assessment (MOCA),^19^ Barthel Index (BI), self-selected gait speed (GS), age, and sex. Participants wore an ActiGraph wGT3X-BT (AG) (https://actigraphcorp.com/actigraph-wgt3x-bt/) 24 hours a day for 5–7 days on their least affected wrist to estimate sleep parameters.^20^ Actigraphs were set to collect data at a 30Hz sample rate and later uploaded for analysis as 60 second epochs with three-axis motion detection. At the conclusion of each timepoint’s wear period, watch data were transferred from the AG to a study computer for analysis using ActiGraph’s ActiLife v6.13.4 software. Wear times were validated (marked as “wear” if either Troiano method or ActiGraph sensor indicated “wear”), and sleep parameters were determined via the Cole Kripke algorithm and Actigraph sleep period detection method. First-pass software-calculations of time-to-bed and time-out-of-bed were sometimes adjusted prior to calculation of sleep parameter variables, under certain conditions. For overnight timeframes with two separate sleep periods detected by the software, a single sleep period spanning the combination of both originally detected periods was substituted. If the ActiLife software was unable to determine time to bed and time out of bed, every effort was made to manually determine these from activity and light signals in the ActiLife interface. Any sleep periods appearing to be daytime naps were not included in analysis of nightly sleep.

### Sleep Health

We developed a 12-point surrogate Ru-SATED (S-Ru-SATED) score adapted from the framework of sleep health proposed by Buysse^11^ that combines sleep parameters (**R**egularity, **T**iming, **E**fficiency, and **D**uration) from actigraphy and self-report of sleep **S**atisfaction and **A**lertness. Regularity was scored based on consistency in bedtimes and wake times, with a score of 2 assigned if deviations were within ±1 hour of the median of bedtime and wake time every night and morning, 1 if this occurred on all but one night or morning, and 0 for two or more deviations. Timing was scored by whether participants slept between 2–4 am, with scores of 2, 1, and 0 assigned for meeting this criterion every night, all but one night, and more than one night, respectively. Efficiency was scored based on nightly sleep efficiency, with 2 assigned if all nights were ≥85%, 1 for all but one night, and 0 for two or more nights <85%. Duration score was based on total sleep time, with a score of 2 for sleeping 7–9 hours each night, 1 for all but one night, and 0 for two or more nights outside this range. Satisfaction was determined from question 4 of the ISI question that asks respondents about their satisfaction with their sleep, with scores of 2, 1, and 0 corresponding to “very satisfied” or “satisfied,” “somewhat dissatisfied,” and “dissatisfied” or “very dissatisfied,” respectively. Alertness was scored using the ESS, with scores of 2 for ESS <11, 1 for scores of 11–12, and 0 for scores >12. The total S-Ru-SATED score ranged from 0 to 12, with higher scores indicating better sleep health. Table 1 summarizes the scoring of the S-Ru-SATED compared to the Ru-SATED self-report measure of SH.

**Table 1.** Surrogate Ru-SATED Scale.

| Item and Definition from Ru-SATED<sup>a</sup> | S-Ru-SATED<sup>b</sup> |  |  | Ru-SATED<sup>a</sup> |  |  |
| --- | --- | --- | --- | --- | --- | --- |
| **Regularity:** | 2: Participant went to bed and got out of bed every night and morning within 1 hour of the median time went to bed and got out of bed. | 1: Participant went to bed and got out of bed all but one night or morning within 1 hour of the median time went to bed and bed and got out of bed. | 0: Participant went to bed and got out of bed all more than one night or morning within 1 hour of the median time went to bed and got out of bed. | 2: Usually/Always | 1: Sometimes | 0: Rarely/Never |
| **Satisfaction:**<br>Are you satisfied | 2: 0 or 1 ISI<sup>c</sup> satisfaction | 1: 2 on ISI<sup>c</sup> satisfaction | 0: 3 or 4 on ISI<sup>c</sup> satisfaction with sleep question | 2: Usually/Always | 1: Sometimes | 0: Rarely/Never |
| with your sleep? | with sleep question | with sleep question |  |  |  |  |
| **Alertness:**<br>Do you stay awake all day without dozing? | 2: <11 on ESS<sup>d</sup> | 1: 11 or 12 on the ESS<sup>d</sup> | 0: >12 on the ESS<sup>d</sup> | 2: Usually/Always | 1: Sometimes | 0: Rarely/Never |
| **Timing:** Are you asleep (or trying to sleep) between 2a and 4a? | 2: Participant was in bed every night between 2a and 4a. | 1: Participant was in bed all but one night between 2a and 4a. | 0: Participant was not in bed more than one night between 2a and 4a. | 2: Usually/Always | 1: Sometimes | 0: Rarely/Never |
| **Efficiency:**<br>Do you spend less than 30 minutes awake at night? | 2: Sleep efficiency 85% or greater every night. | 1: Sleep efficiency 85% or greater all but one night. | 0: Sleep efficiency less than 85% two or more nights. | 2: Usually/Always | 1: Sometimes | 0: Rarely/Never |
| **Duration:**<br>Do you sleep between 6 | 2: Total sleep time between 7 | 1: Total sleep time between 7 and 9 hours | 2: More than one night during which total | 2: Usually/Always | 1: Sometimes | 0: Rarely/Never |
| and 8 hours<br>per day? | and 9 hours<br>each night. | for all but<br>one night. | sleep time was<br>not between 7<br>and 9 hours. |  |  |  |

<sup>a</sup>Ru-SATED, Regularity, Satisfaction, Alertness, Timing, Efficiency, Duration; <sup>b</sup>S-Ru-SATED, Surrogate Ru-SATED; <sup>c</sup>ISI, Insomnia Severity Index; <sup>d</sup>ESS, Epworth Sleepiness Scale.

### Data Analysis

Descriptive statistics were used to summarize the sample characteristics. To investigate factors influencing SH, we employed a cumulative link mixed model (CLMM), a robust method for analyzing ordinal response data. Specifically, the CLMM approach estimates the probability that the response (outcome-SH) is at or below that category (the cumulative probability that response *Y* is less than or equal to some category *k*). The logit-transformed cumulative probability is written as a function of fixed effects defined as predictive covariates, including PHQ-9 (depression), ODI, MOCA (cognitive function), BI (functional ability), GS (walking capability), age, sex, and subject-level random intercepts to account for the repeated measures across time. The mathematical representation of the proposed model is given by: S- Ru-SATED ∼ Sex + PHQ-9 + BI + age + ODI + GS + MOCA + Time post stroke + (1|participant). Before developing the model, we examined the correlation among the covariates to not include covariates that were associated with each other in the model to avoid multicollinearity.

## Results

Data from 90 participants was used in the analyses and collected at a mean (standard deviation) 9.4 (7.5), 60.0 (4), and 94.0 (6.0) days post stroke. Participants were a mean (standard deviation) of 64.3 (15.8) years old, 57% (n=51) were female, 74% (n=67) identified as white, 23% (n=21) identified as Black, and 2% (n=2) identified as more than one race, see Table 2.

**Table 2.**
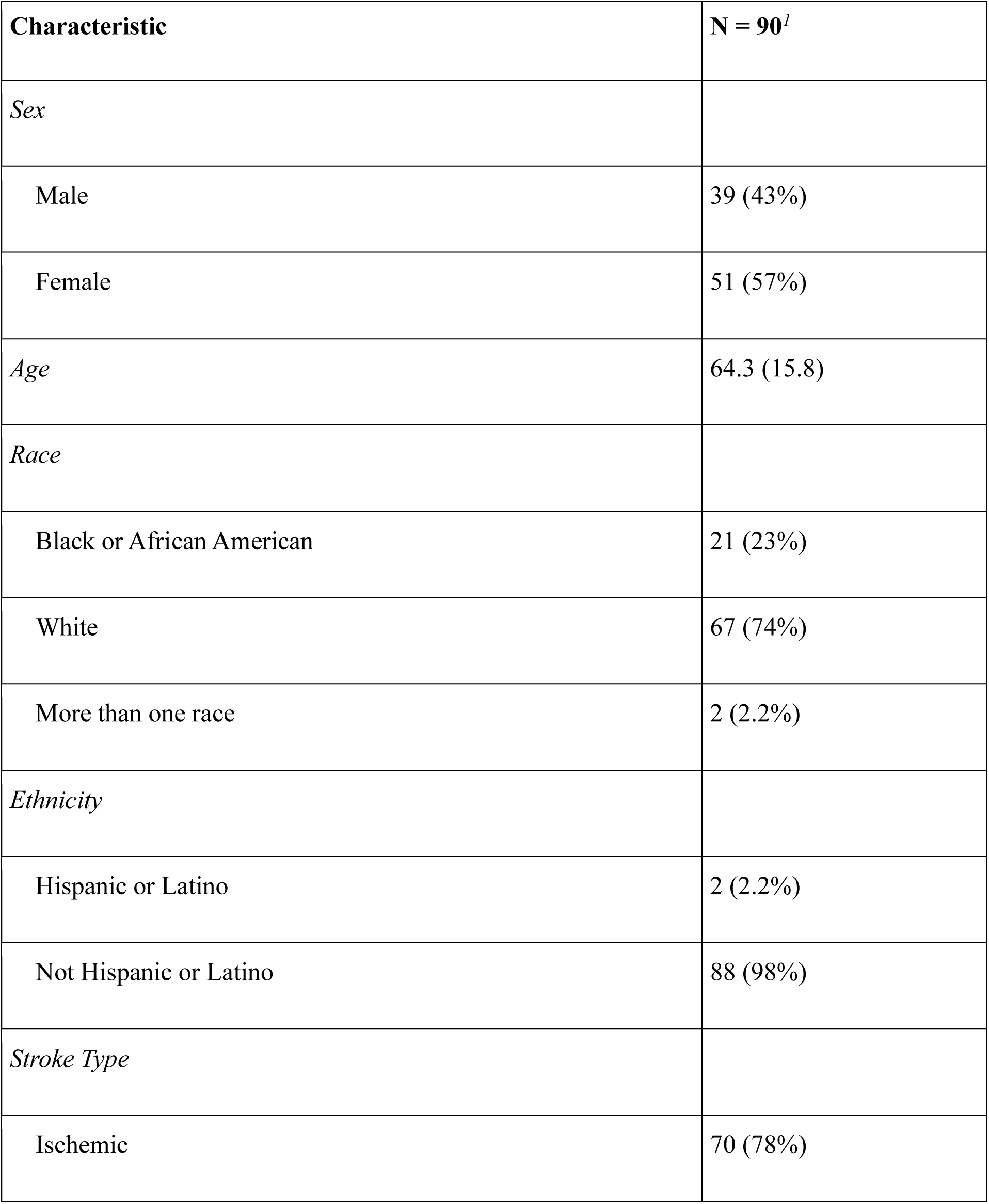
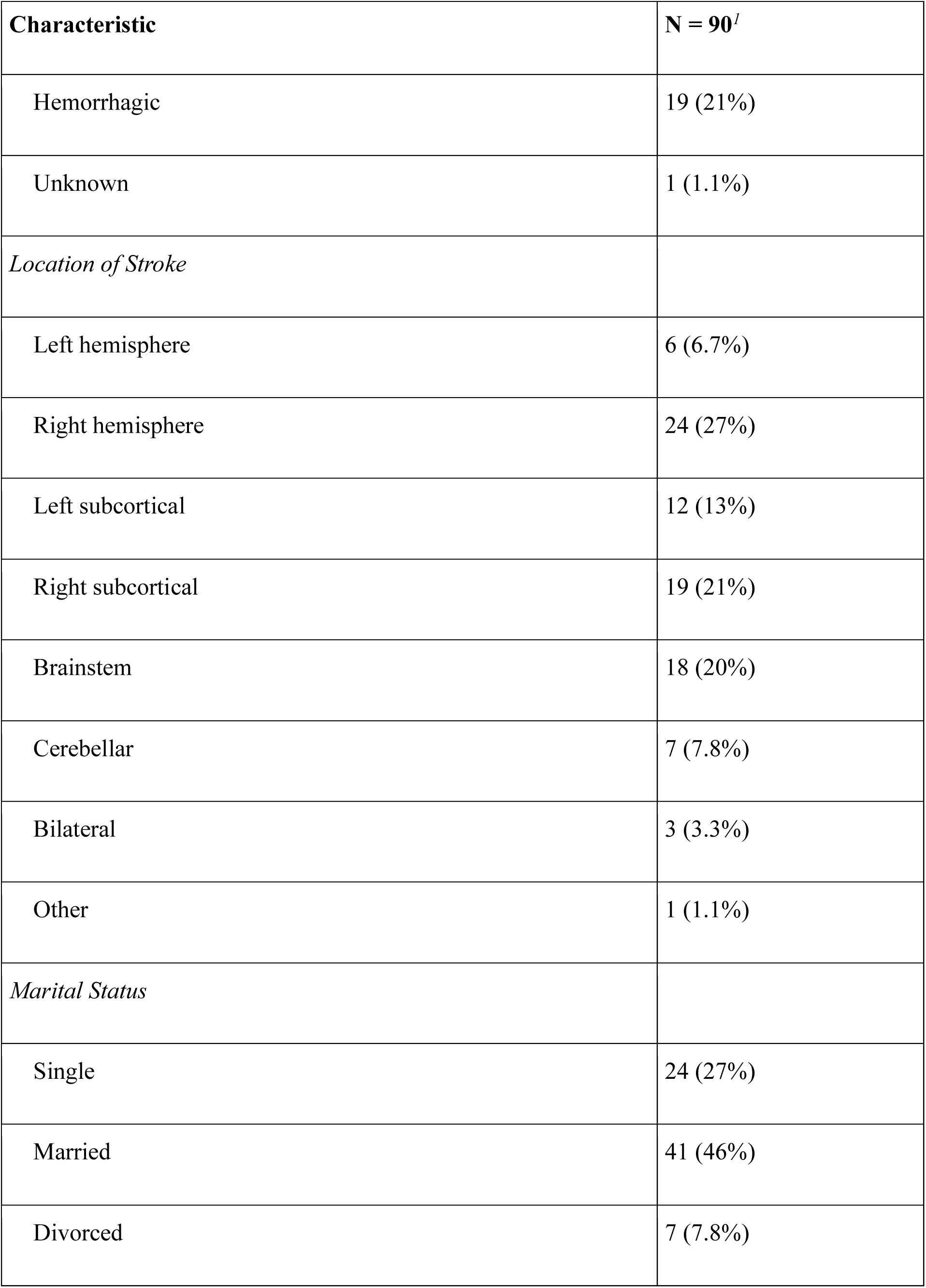
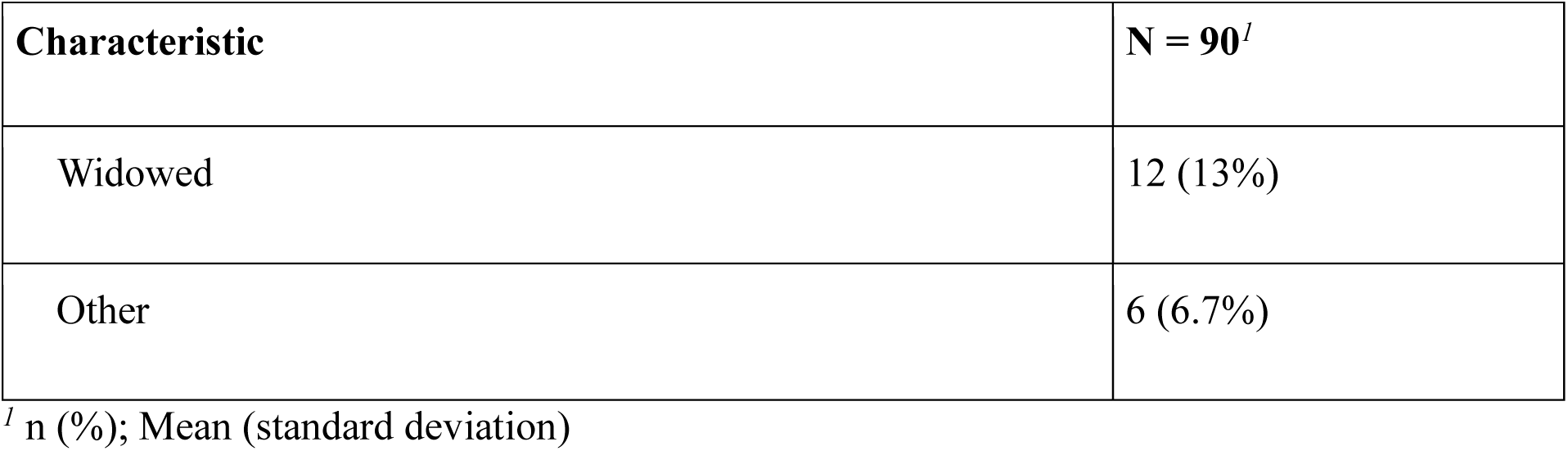
Participant Characteristics.

| Characteristic | N = 90<sup>a</sup> |
| --- | --- |
| *Sex* |  |
| Male | 39 (43%) |
| Female | 51 (57%) |
| *Age* | 64.3 (15.8) |
| *Race* |  |
| Black or African American | 21 (23%) |
| White | 67 (74%) |
| More than one race | 2 (2.2%) |
| *Ethnicity* |  |
| Hispanic or Latino | 2 (2.2%) |
| Not Hispanic or Latino | 88 (98%) |
| *Stroke Type* |  |
| Ischemic | 70 (78%) |

| **Characteristic** | **N = 90<sup>a</sup>** |
| --- | --- |
| Hemorrhagic | 19 (21%) |
| Unknown | 1 (1.1%) |
| *Location of Stroke* |  |
| Left hemisphere | 6 (6.7%) |
| Right hemisphere | 24 (27%) |
| Left subcortical | 12 (13%) |
| Right subcortical | 19 (21%) |
| Brainstem | 18 (20%) |
| Cerebellar | 7 (7.8%) |
| Bilateral | 3 (3.3%) |
| Other | 1 (1.1%) |
| *Marital Status* |  |
| Single | 24 (27%) |
| Married | 41 (46%) |
| Divorced | 7 (7.8%) |

| Characteristic | N = 90<sup>1</sup> |
| --- | --- |
| Widowed | 12 (13%) |
| Other | 6 (6.7%) |

<sup>1</sup> n (%); Mean (standard deviation)

Participants wore the AG for a mean of 4.2 nights at 10 days post stroke during inpatient rehabilitation, 6.7 nights at 60 days post stroke, and 6.4 nights at 90 days post stroke. See Table 3 and Figure 1 for S-Ru-SATED scores at 10-, 60-, and 90- days post stroke.

**Figure 1.**
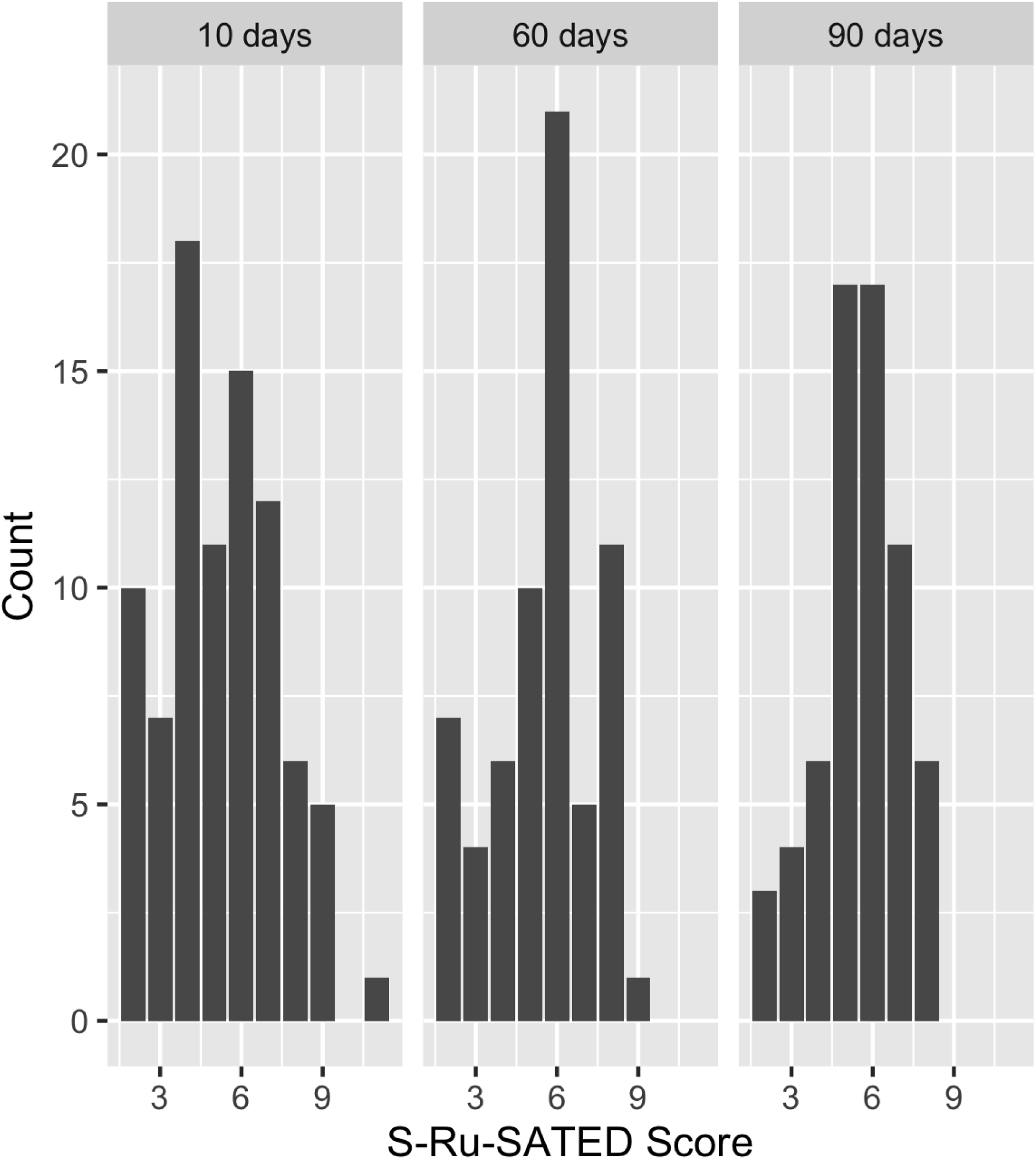
Surrogate Ru-SATED Scores During Recovery After Stroke

**Table 3.** S-Ru-SATED, Sleep Parameters, and Co-Variates at 10-, 60-, and 90-days Post Stroke.

| Characteristic | 10 days post stroke | 60 days post stroke | 90 days post stroke |
| --- | --- | --- | --- |
| S-Ru-SATED,<sup>a</sup> median (range) | 5.0 (2.0-10.0) | 6.0 (2.0-9.0) | 6.0 (2.0-8.0) |
| Time into Bed, hh:mm,<sup>b</sup> median | 18:25 | 18:57 | 17:38 |
| Time out of Bed, hh:mm, median | 06:13 | 07:39 | 08:10 |
| ISI<sup>c</sup> satisfaction with sleep question, median (range) | 2 (0-4) | 1 (0-4) | 2 (0-4) |
| ESS,<sup>d</sup> mean (sd)<sup>e</sup> | 8.9 (5.4) | 6.9 (5.0) | 5.6 (4.1) |
| Sleep Efficiency, %, mean (sd) | 83.19 (9.49) | 82.32 (8.80) | 81.24 (9.25) |
| Duration of sleep, minutes, mean (sd) | 411.63 (100.96) | 463.66 (88.71) | 453.32 (100.91) |
| Oxygen desaturation index, mean (sd) | 5.69 (4.06) | 8.39 (7.06) | 9.14 (8.92) |
| Barthel Index, mean (sd) | 42.24 (22.26) | 85.31 (20.25) | 91.17 (14.87) |
| Self-selected Gait Speed, mean (sd) | 0.23 (0.29) | 0.61 (0.41) | 0.70 (0.39) |
| Patient Health Questionnaire-9, mean (sd) | 4.89 (5.30) | 4.45 (4.45) | 3.83 (4.47) |
| Montreal Cognitive Assessment, mean (sd) | 21.22 (5.44) | 23.22 (4.47) | 24.40 (4.53) |

<sup>a</sup>S-Ru-SATED, Surrogate Ru-SATED; <sup>b</sup>hh:mm, hour:hour:minute:minute; <sup>c</sup>ISI, Insomnia
Severity Scale; <sup>d</sup>ESS, Epworth Sleepiness Scale; <sup>e</sup>sd, standard deviation

There were significant fair to strong associations between BI and GS (r=0.77) and BI and MOCA (r=0.35) as a result we did not include GS or MOCA as independent variables in the CLMM model to avoid spurious trends due to multicollinearity. The CLMM found the fixed effects of the PHQ-9 had a significant negative estimate of-0.116 (p = 0.0002), indicating that for each unit increase in PHQ-9, the odds of having a one unit increase in the S-RU-SATED score decreased by approximately 11% holding other variables constant. This means that higher levels of depressive symptoms are associated with a reduced likelihood of good sleep health. The BI had a significant estimate of 0.016 (p = 0.044), indicating that for each unit increase in the BI, the odds of having a one unit increase in the S-Ru-SATED score increased by approximately 1.6% holding other variables constant. This indicates that greater functional independence is associated with slightly higher odds of better sleep health. The other predictors were not statistically significant. Sex had an estimate of-0.130 (p = 0.650), indicating no meaningful difference in SH between females and males. Age had a positive estimate of 0.0150 (p = 0.116), suggesting a weak and non-significant trend where older individuals might have slightly higher odds of having better SH. Time post stroke showed non-significant negative estimates suggesting no substantial change in SH between 10-, 60-, and 90- days post stroke. Subject-specific random effects revealed a variance of 0.1714 and a standard deviation of 0.4139, indicating large variability in the intercept across groups.

The threshold coefficients represent the log-odds separating adjacent S-Ru-SATED scores, indicating the difficulty of transitioning between them. Lower thresholds, such as 2∣3 (-2.14), suggest that improving from a score of 2 to a score of 3 is relatively easier, while higher thresholds, like 9∣11 (6.26), suggest that achieving the highest sleep health score is substantially more difficult, see Table 4.

**Table 4.** S-Ru-SATED Threshold Coefficients.

| Threshold | Estimate | Std. Error |
| --- | --- | --- |
| 2\|3 | -2.1387 | 0.7151 |
| 3\|4 | -1.2695 | 0.6858 |
| 4\|5 | -0.2135 | 0.6685 |
| 5\|6 | 0.6894 | 0.6643 |
| 6\|7 | 1.9285 | 0.6777 |
| 7\|8 | 2.8828 | 0.7068 |
| 8\|9 | 4.6243 | 0.8261 |

## Discussion

In our sample of people after stroke without moderate to severe obstructive sleep apnea (OSA), we identified depression and functional ability as significant determinants of SH. Higher levels of depressive symptoms, as assessed by the PHQ-9, and diminished functional ability, as measured by the BI, were strongly associated with poorer SH. Notably, time since stroke did not influence SH, as participants exhibited persistently poor SH that remained unchanged after discharge. These findings illustrate the complex, bidirectional nature of sleep health and recovery after stroke.

Sleep is impacted by multiple factors after stroke and can influence outcomes after stroke. Disturbed sleep may lead to the development of specific sleep disorders, which in turn can lead to poorer outcomes.^3^ Our study is unique in that we examined sleep from a broader perspective of SH rather than individual sleep parameters or specific sleep disorders, and we did not include people with possible moderate to severe OSA. The results highlight the importance of SH for all individuals post stroke, not just those with sleep-disordered breathing.

Depression is common and can be persistent after stroke.^21^ As we hypothesized, our findings suggest that depression may contribute to poor SH during recovery after stroke. Participants with higher levels of depression had greater odds of poor SH. This is supported by findings from other authors. Fan and colleagues^5^ reported that people with poor sleep quality, measured by the Pittsburgh Sleep Quality Index (PSQI), at two weeks post stroke that persisted at three months had an increased risk of depression. In a cross-sectional analysis of people with chronic stroke, Silva and colleagues^6^ found that depression explained 22% of the variance in sleep quality, measured by the PSQI.

People with stroke with sleep disorders exhibit poorer functional ability than those without sleep disorders.^3^ As hypothesized, we found that poorer functional ability was associated with poorer SH. Similarly, Silva and colleagues^6^ reported that depression combined with disability explained 30% of the variance in sleep quality. Denis and colleagues^9^ found that decreased total sleep time at 15 days after stroke was associated with greater disability for people at 3 months post stroke. However, Fan and colleagues^5^ found that poor sleep health at two weeks after stroke that persisted at three months was not associated with poorer function. Unlike these studies, and those reported on in the preceding paragraph, we used both objective and subjective measures to create a composite measure of SH rather than a single subjective measure of sleep quality, which further highlights the impact of depression and function on sleep after stroke. Cognitive behavioral therapy for insomnia (CBT-I) is an effective treatment to improve sleep in people with depression with insomnia and may also improve depression symptoms.^22^ Given the influence of depression on SH in people with stroke, a behavioral sleep health intervention may be helpful and should be explored in people with stroke.

Although we hypothesized that walking capability (measured by GS) and cognition (measured by the MOCA) would also significantly influence SH, our findings did not support this. Li and colleagues^10^ found that people early after mild stroke who demonstrated cognitive impairment had less total sleep time than those without cognitive impairment. Unlike our study, Li and colleagues used comprehensive battery of tests to assess cognitive impairment. While we used the MOCA, which is a screening exam of global cognition. Because GS is a strong measure of walking capability, which is associated with activity^23^ we hypothesized that it would also influence SH. However, due to its association with the BI, which is a broader measure of functional ability, it was not included in our model.

The finding that time post stroke did not influence SH was unexpected. Sleep health was poor at 10-, 60-, and 90-days post stroke and did not significantly change after participants left the hospital. Burger and colleagues^24^ found that 76% of people who were hospitalized had poor sleep quality and quantity. Hospitalized adults experienced 1.3-3.2 hours less than is recommended.

Wesselius and colleagues^25^ reported that adults experienced 83 minutes less sleep/night in the hospital compared to at home and that their sleep quality was significantly worse in the hospital. One possible explanation for this difference is that we measured SH, not specific sleep parameters or sleep quality and that individual items on the S-Ru-SATED are based on consistency over approximately one week (the duration of time that participants wore the AG) and not an average of sleep parameters. Consistency of sleep may be an important aspect of SH to consider.

A limitation of our study is that we did not include people with stroke who have moderate to severe OSA in our sample, so the results need to be interpreted with this in mind. Obstructive sleep apnea is common in people with stroke and people with OSA likely have poor SH. Further research that examines SH in this group of people after stroke is needed. There may be factors in addition to depression and functional ability that impact SH in people with stroke with OSA. Another limitation is that we did not use the Ru-SATED to measure SH, which is commonly used to measure SH.^11^ The S-Ru-SATED developed and used in this study is an aggregate measure of SH that combined objective and subjective sleep parameters identified as key components of SH. We also do not know the pre-stroke sleep health and depression of the participants, which may have impacted their recovery post-stroke.

## Conclusion

Depression and functional ability are associated with SH during recovery in people with stroke without moderate to severe OSA. Greater symptoms of depression and lower functional ability are associated with poorer SH. Sleep health was poor early after stroke during inpatient rehabilitation and did not improve when participants were back in their homes. Future research is needed to examine the effect of improving SH on recovery after stroke.

## Data Availability

Data are not available for this study at the present time.

